# An Assessment of Correctional Officers’ Health Beliefs in Relationship to COVID-19 Vaccine Uptake and Hesitancy

**DOI:** 10.64898/2026.03.24.26349101

**Authors:** Bethany Hedden-Clayton, Ariel L. Roddy, Juliette K. Roddy, Yvane Ngassa, Bridget Pickard, Rachel A. Tam, Alysse G. Wurcel

## Abstract

**Introduction:** During the COVID-19 pandemic, incarcerated populations faced heightened risk of exposure due to healthcare barriers, restrictive environments, and pre-existing health conditions. Consequently, Correctional Officers (COs) faced increased risk of COVID-19 exposure. Given the health benefits of COVID-19 vaccination and the rise in vaccine hesitancy, this study examined the relationship between COs health beliefs and COVID-19 vaccine uptake.

**Methods:** A health beliefs survey was administered to Massachusetts-based COs (n=118). Chi-squared Automatic Interaction Detection modeling and logistic regression was utilized to analyze the survey data.

**Results:** COs with higher trust in vaccines and a prior positive COVID-19 test were most likely to get vaccinated voluntarily. Those with low trust in vaccines and no previous positive COVID-19 test were least likely to receive the vaccine.

**Conclusion:** Despite the severe impact of COVID-19 in correctional settings, and the evidence of vaccine efficacy against hospitalization and death, vaccine uptake among COs remains low.

## Introduction

Since the emergence of the global SARS-CoV-2 (COVID-19) pandemic in December 2019, vaccine hesitancy and the anti-vaccine (anti-vax) movement have gained increased visibility. As of this writing, there have been over 7 million reported COVID-19 deaths worldwide(World Health Organization, 2023) and over 1 million deaths in the U.S. alone (Ahmad FB, 2025). While personal protective equipment and behaviors – face masks, hand-washing, social distancing – are helpful in slowing the spread of COVID-19, the most effective tool is vaccination (Nandi & Shet, 2020). Even though the COVID-19 vaccine was widely available in the U.S. by April 2021 (Diesel J, 2021), only 79% of adults had completed the primary COVID-19 vaccination series. During the era of COVID-19 and after the vaccine was developed, vaccine uptake was hindered by vaccine hesitancy. The World Health Organization’s SAGE (Strategic Advisory Group of Experts on Immunization) defines vaccine hesitancy as the refusal of vaccination despite its availability (MacDonald, 2015). Recent studies show that approximately 40% of Americans are concerned about or reject the COVID-19 vaccine (Hyatt et al., 2021).

Vaccine hesitancy is particularly important to address within essential workers. The “essential workers” category is heterogeneous and has varying levels of exposure to COVID-19. While many of the essential workforce are healthcare workers, service workers, transportation, and law enforcement employees are also included. Vaccine hesitancy was approximately 7% for essential workers within healthcare, however it was 14% for non-essential workers, and 17% for essential workers outside of healthcare (Beleche et al., 2023). A subgroup within the essential and non-healthcare workers are Correctional Officers (COs).

COs face unique occupational risks highlighted during the COVID-19 pandemic. The work COs do puts them at an increased risk of contracting an infectious disease like COVID-19. COs work in jails, prisons, and other carceral settings and supervise people in custody (U.S. Department of Labor, 2025). These facilities are often overcrowded, poorly ventilated, and unsanitary, with frequent population turnover, which increases the risk of infectious disease exposure (Kinner et al., 2020). Additionally, carceral settings concentrate populations who are more susceptible to infections and COVID-19-related complications (Kinner et al., 2020). As a result, COs are uniquely vulnerable to COVID-19. By August 2020, COVID-19 cases were 5 times higher within carceral settings compared with the general U.S. population, affecting not only people who are incarcerated but also correctional staff and the communities to which individuals return (National Academies of Sciences et al., 2020). The pandemic revealed both the limited capacity of carceral healthcare systems and the interconnectedness of correctional facilities with the general population (Kinner et al., 2020). When people are released from jail or prison, they can potentially spread COVID-19 and disproportionately impact marginalized communities (Kinner et al., 2020). In order to mitigate the high rates of infection in carceral settings in both people who are incarcerated and people who are employed by carceral systems, it is important to address the vaccine hesitancy within COs.

Due to the scant literature about COVID-19 vaccine uptake within COs, we looked at vaccine hesitancy in the broader group of law enforcement officers and first responders (Apprenticeship USA, 2025). Previous studies examining COVID-19 vaccination attitudes among law enforcement officers and first responders have consistently highlighted a theme of mistrust (Caban-Martinez et al., 2022). Mistrust is the general sense of suspicion towards someone or something from the belief that they will not act in that person’s best interest (Griffith et al., 2021). A similar yet distinct concept is that of distrust. Distrust is based on personal or collective prior experiences that lead to increased skepticism and suspicion (Griffith et al., 2021). In summary, mistrust is the lack of faith in something because of a suspicion, and distrust is the lack of faith based on knowledge or prior experiences. Participants reported mistrust of the government, media sources, and doctors (Caban-Martinez et al., 2022). Building on this, we aimed to investigate the relationship between COVID-19 vaccine uptake and general health beliefs among COs, to determine whether vaccine hesitancy was associated with other non-evidence-based health beliefs. The survey we administered included questions on vaccine attitudes, chronic illness, emergency preparedness, behavioral health, political influences, and personal experiences with the COVID-19 pandemic. This research seeks to elucidate how health beliefs shape COs’ perspectives on current and future vaccination efforts.

## Study Methods

### Current Study

The current study aimed to examine the relationship between COs’ health beliefs and their COVID-19 vaccine uptake. Survey data was collected from jail- and prison-based COs located in Massachusetts (N=118) to investigate how an array of health beliefs related to COVID-19 vaccine receipt. This human subjects research project was assessed by the Health Sciences Institutional Review Board at Tufts Medical Center. Approval was granted in May 2022. Health beliefs assessed included attitudes toward vaccines, perceptions of chronic illness, endorsement of emergency preparedness behaviors, perspectives on behavioral health treatments, and attitudes toward public health policies, regulations, and government intervention. Responses were aggregated into scales, which were then analyzed in relation to COVID-19 vaccine uptake using Chi-squared Automatic Interaction Detection (CHAID) modeling.

### Sample

We employed convenience sampling to administer an anonymous 49-item health beliefs survey at in-person correction officer union meetings and jails in a New England state. The survey, developed by the authors, consisted of three sections: health-related beliefs and attitudes, personal experiences with COVID-19, and brief demographic information (see Supplemental Material for survey instrument). The data underlying this article will be shared on reasonable request to the corresponding author. Two members of the research team conducted survey administration between May-August 2022. Participants received a $50 gift certificate to a home improvement retailer for completing the survey.

### Independent Variables

The survey instrument was used to construct five distinct scales, which served as the independent variables in this study. Each scale assessed a different aspect of health-related beliefs and attitudes and consisted of five items, with responses recorded on a Likert scale ranging from Strongly Agree (=3) to Strongly Disagree (=0).

#### Vaccine Attitudes

This scale measured perceptions of vaccine safety and necessity. The following five items were included: “Vaccines are safe,” “The Hepatitis B vaccine is safe,” “The immune system can keep the body healthy without medications or vaccines” (reverse coded), “The COVID-19 vaccines are safe,” and “Mandated vaccination against COVID-19 is necessary for public health.” Higher scores reflected greater trust in vaccines, whereas lower scores indicated vaccine hesitancy or opposition. Responses were measured on a 4-point Likert scale, where 0 = “Strongly disagree” and 3 = “Strongly agree.”

#### Chronic Illness

This scale measured respondents’ confidence in established medical treatments and preventative measures. The following five items were included: “Sunblock helps to prevent skin cancer,” “Colonoscopies can help to prevent colon cancer,” “Insulin is necessary to treat people with diabetes,” “Smoking cigarettes is healthy” (reverse coded), and “High blood pressure can be treated with medications.” Higher scores reflected stronger confidence in evidence-based medical interventions, whereas lower scores indicated skepticism or uncertainty.

#### Emergency Preparedness

This scale evaluated attitudes toward emergency preparedness, safety precautions and risk-mitigation behaviors. It included five items: “Seatbelts can save lives,” “Fire detectors are a proven way to prevent death,” “It is important to have homeowners or renters insurance for your home,” “Helmets should be worn when riding a motorcycle,” and “It is important to know where to shelter in case of an emergency.” Higher scores reflected greater endorsement of safety-related behaviors and protective measures.

#### Behavioral Health

This scale measured perceptions of behavioral health, mental well-being, and substance use treatment. Items included: “Medication can treat depression,” “Vaccines cause mental illness” (reverse coded), “Overuse of caffeine can disrupt sleep,” “Meditation can help reduce stress,” and “There are effective treatments for substance use disorder.” Higher scores indicated stronger acceptance of evidence-based behavioral health concepts, whereas lower scores suggested skepticism or misinformation.

#### Political Intersections

This scale captured attitudes toward public health policies, regulations, and government intervention. Items included: “Businesses should have the right to turn away customers based on vaccination status,” “Public health benefits gained from government-imposed restrictions on businesses during the pandemic outweigh the costs,” “Public health officials are hiding things they know about the COVID-19 vaccine” (reverse coded), “To promote public health, states should impose a higher tax on soda and candy bars,” and “A high public vaccination rate is necessary for reducing communicable diseases such as measles, mumps, polio, and rubella.” Higher scores reflected stronger support for public health initiatives, whereas lower scores indicated skepticism or opposition.

#### Personal Experiences with COVID-19

The second section of the survey included 17 binary questions (Yes/No) addressing respondents’ experiences with COVID-19. Seven items assessed self-reported adherence to COVID-19 mitigation behaviors such as social distancing, use of personal protective equipment, hand washing and sanitizing, and vaccination against COVID-19. Six items focused on personal experiences with COVID-19 exposure and risk, such as having been tested for COVID-19, receiving a positive test result, or being hospitalized for COVID-19. Respondents were also asked whether they personally knew someone who had tested positive or had been hospitalized for COVID-19. Two additional items evaluated respondents’ acceptability of the state’s public health response to the pandemic.

#### Demographic Information Items

The final section of the survey included two demographic variables – highest level of education and age – which were used as control variables. Education level was categorized into five groups: “High school or GED” (46.74%), “Associates” (19.57%), “Bachelors” (28.26%), “Masters” (4.35%), and “PhD” (1.09%). Each category was coded as a binary variable (0 = No, 1 = Yes). Age was grouped into five generational cohorts: “1928-1945” (1.09%), “1946-1964” (8.70%), “1965-1980” (54.35%), “1981-1995” (35.87%), and “1996-2010” (1.09%), which correspond to Traditionalists, Baby Boomers, Generation X, Millennials, and Generation Z, respectively. Each cohort was coded as a binary variable (0 = No, 1 = Yes). Based on the authors’ practice-experience and existing relationships with participants in this state, demographic items on race/ethnicity and gender were intentionally excluded, as such questions are often left unanswered and, at worst, could compromise researcher-participant rapport.

### Dependent Variable

The dependent variable was self-reported COVID-19 vaccination status, coded as 1 for individuals who reported receiving the vaccine and 0 for those who did not.

### Analytic Approach

Before conducting formal analyses, we examined summary statistics for all dependent and independent variables. The analysis proceeded in two stages.

#### Stage 1: Decision Tree Modeling

In the first stage, we used decision tree modeling to identify the characteristics associated with COVID-19 uptake among COs. Traditional parametric regression techniques have several limitations, including sensitivity to multicollinearity, the need to pre-specify interactions and functional relationships between variables, and poor handling of missing data. To address these challenges, we employed Chi-squared Automatic Interaction Detection modeling (Kass, 1980), an alternative approach for predicting categorical response variables when relationships among predictors are unspecified and theoretical guidance is limited.

CHAID modeling iteratively splits the sample into mutually exclusive (nodes) using predictor variables that best differentiate the outcome of interest. At each step, all possible category pairs across predictor variables are tested, and the variable demonstrating the strongest association with the dependent variable is selected to generate a new node. This process continues until no additional statistically significant splits can be made, resulting in terminal nodes. The top, or root, node represents the full sample, and each successive layer of nodes reflects increasingly specific subgroups. In the resulting classification tree, ovals denote internal nodes and squares denote terminal nodes. CHAID makes no assumptions about underlying data distribution, minimizes the effects of outliers, and accommodates both categorical and ordinal data.

#### Stage 2: Logistic Regression

In the second stage, logistic regression was performed using the dependent variable — COVID-19 vaccine uptake — and the independent variables identified as significant predictors from the CHAID model, along with relevant controls. The logistic regression produced odds ratios representing the likelihood of vaccine uptake associated with a one-unit increase in each independent variable. All analyses were conducted using Stata version 15 (StataCorp, 2017).

### Missing Data

Of the 116 individuals in the sample, 24 participants (20.69%) had more than five missing responses across key variables, potentially compromising the validity of machine learning analyses.(Emmanuel et al., 2021) Missingness appeared to stem from participants’ discomfort in answering certain items, as reflected in skipped questions or written notes in the survey margins. To minimize bias, listwise deletion was applied to cases with more than five missing responses. This resulted in n=92 surveys being included in the analysis.

## Results

### Descriptive Statistics for Key Variables

Table 1 presents the descriptive statistics for the five measured scales, including mean scores, standard deviations, internal consistency (α, Cronbach’s alpha), and observed minimum and maximum values. The Vaccine Attitudes scale had a mean score of 1.62 (SD = 0.71), with responses ranging from 0 to 3. The internal consistency for this scale was α = .75, indicating acceptable reliability. The Chronic Illness scale had a mean score of 2.37 (SD = 0.42), with a minimum observed score of 0 and a maximum of 3. The internal consistency was α = .61, suggesting moderate reliability. The Emergency Preparedness scale had a mean of 2.38 (SD = 0.36) and an internal consistency of α = .58. Responses ranged from 0 to 3, indicating a relatively high overall endorsement of emergency preparedness beliefs. The Behavioral Health scale yielded a mean of 2.03 (SD = 0.43), with responses spanning the full range (0 to 3). However, the internal consistency was lower (α = .51), suggesting a more variable relationship among the included items. Finally, the Political Intersections scale had the lowest mean score (1.33, SD = 0.52) and the lowest internal consistency (α = .49), with responses ranging from 0 to 3, suggesting broader variability and potential measurement limitations. Overall, internal consistency varied across scales, with some scales (e.g., Vaccine Attitudes) demonstrating stronger reliability and others (e.g., Political Intersections, Behavioral Health) reflecting weaker internal consistency.

**Table 1.**
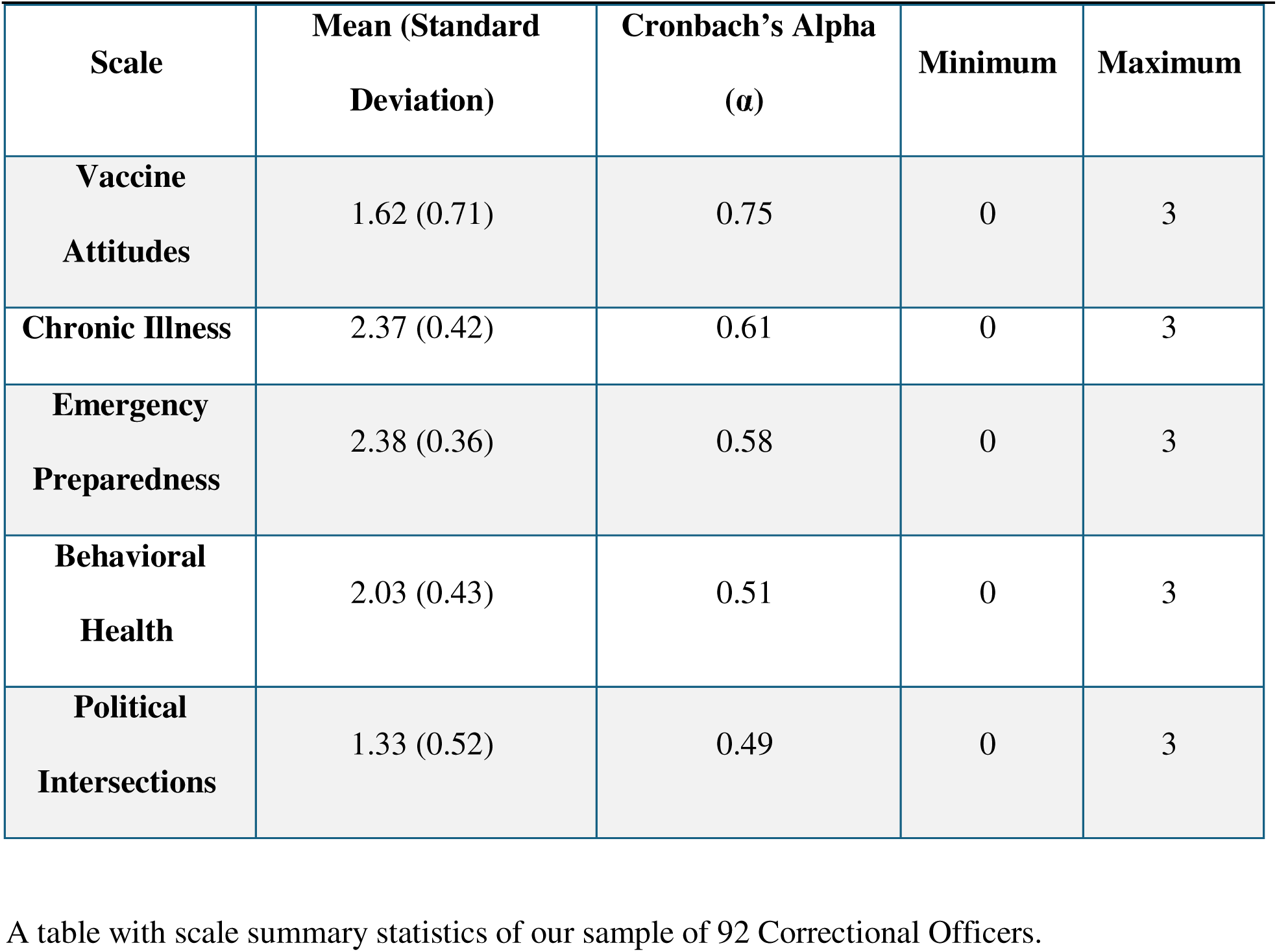
Scale summary statistics (n=92)

Table 2 presents the descriptive statistics of the non-scale variables. Of the total sample, 62 participants (67.39%) reported having received the COVID-19 vaccine, and 39 (42.86%) reported ever testing positive for COVID-19. The majority of respondents had a high school education or equivalent (46.74%, n=43) which is the typical entry-level education requirement (U.S. Department of Labor, 2025), followed by those with an associate’s degree (19.57%, n=18), a bachelor’s degree (28.26%, n=26), and a master’s degree (4.35%, n=4). Only one participant (1.09%) reported holding a PhD. Most participants were born between 1965-1980 (54.35%, n=50) or 1981-1995 (35.87%, n=33). Smaller proportions were born between 1946-1964 (8.70%, n=8), with very few in the 1928-1945 (1.09%, n=1) and 1996-2010 (1.09%, n=1) cohorts.

**Table 2.**
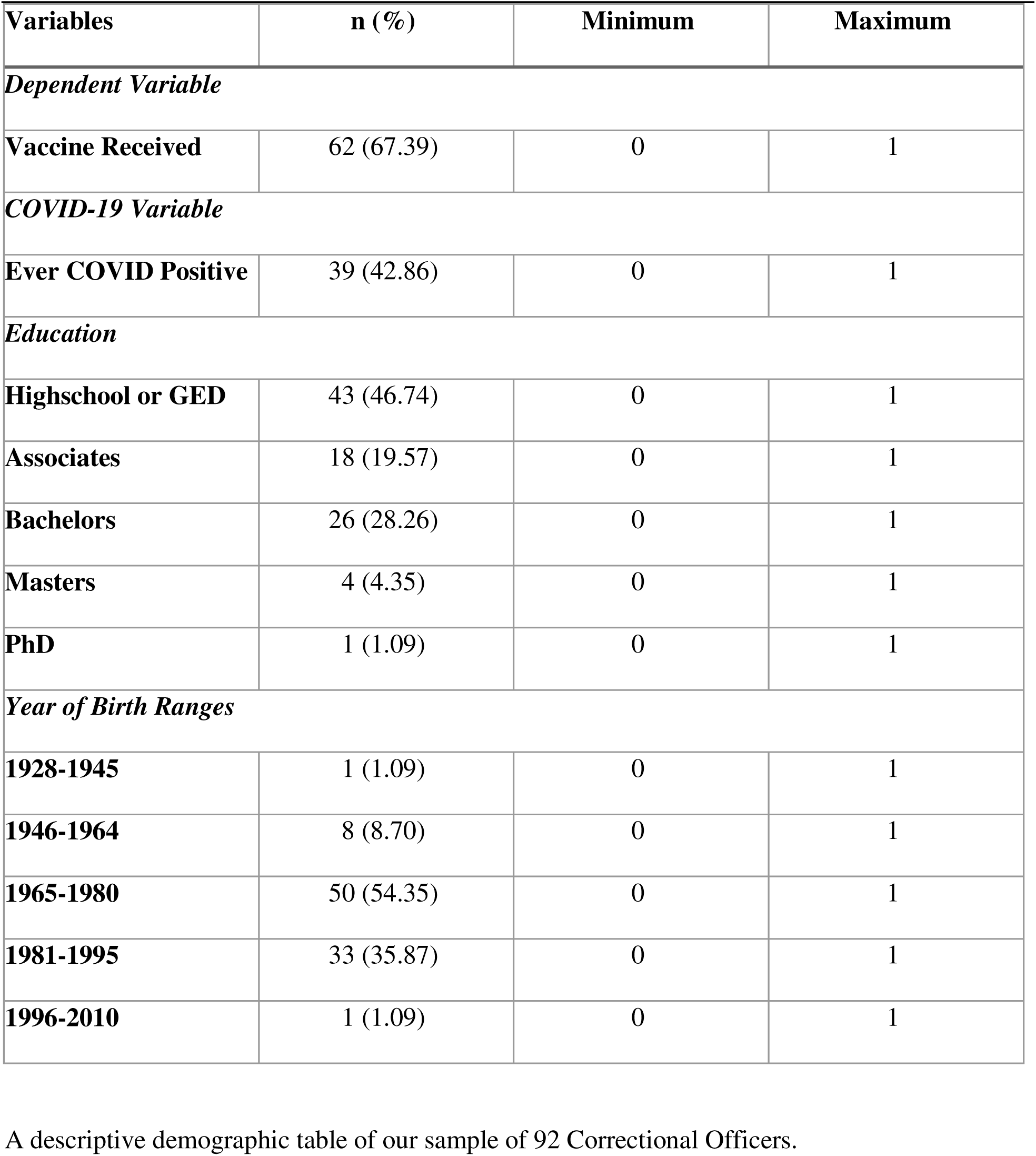
Descriptive statistics for non-scale variables (n=92).

| **Variables** | **n (%)** | **Minimum** | **Maximum** |
| --- | --- | --- | --- |
| ***Dependent Variable*** |  |  |  |
| **Vaccine Received** | 62 (67.39) | 0 | 1 |
| ***COVID-19 Variable*** |  |  |  |
| **Ever COVID Positive** | 39 (42.86) | 0 | 1 |
| ***Education*** |  |  |  |
| **Highschool or GED** | 43 (46.74) | 0 | 1 |
| **Associates** | 18 (19.57) | 0 | 1 |
| **Bachelors** | 26 (28.26) | 0 | 1 |
| **Masters** | 4 (4.35) | 0 | 1 |
| **PhD** | 1 (1.09) | 0 | 1 |
| ***Year of Birth Ranges*** |  |  |  |
| **1928-1945** | 1 (1.09) | 0 | 1 |
| **1946-1964** | 8 (8.70) | 0 | 1 |
| **1965-1980** | 50 (54.35) | 0 | 1 |
| **1981-1995** | 33 (35.87) | 0 | 1 |
| **1996-2010** | 1 (1.09) | 0 | 1 |

A descriptive demographic table of our sample of 92 Correctional Officers.

### Model 1: CHAID Analysis Results

Figure 1 presents the results of the CHAID decision tree model predicting COVID-19 vaccine uptake for COs. Two variables, Vaccine Attitudes scale and COVID-19 infection history, emerged as the strongest predictors of vaccine uptake. No other scale or control variables were selected by the model.

**Figure 1.**
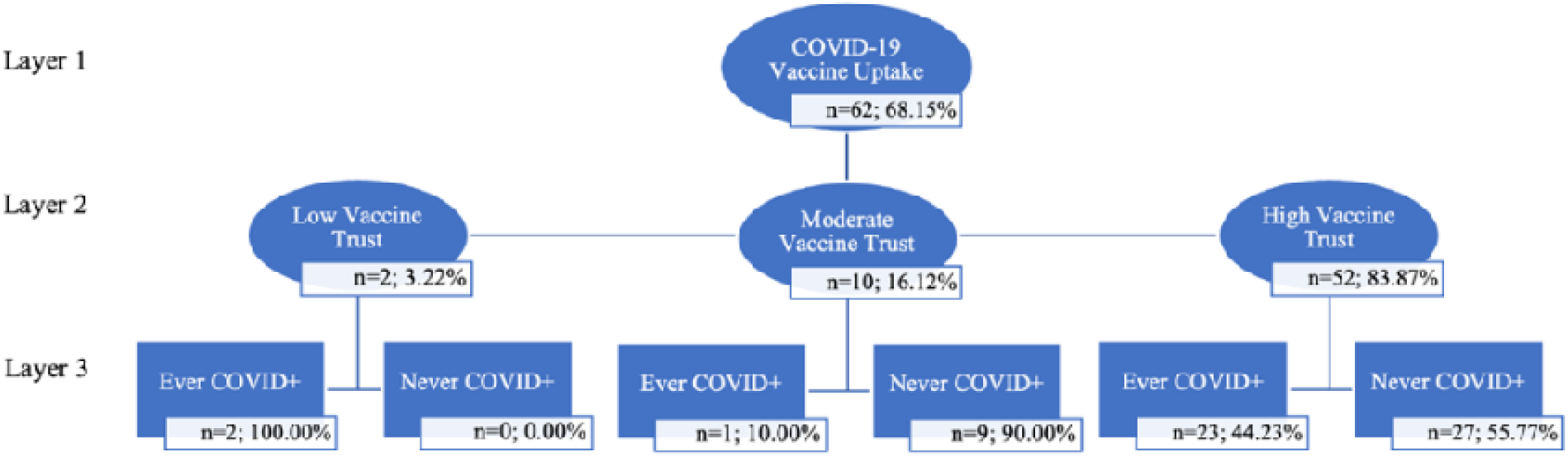
Decision tree of characteristics predicting COVID-19 vaccine uptake among Correctional Officers. A decision tree generated using Chi-square Automatic Interaction Detection (CHAID) modeling. Ovals represent internal nodes and squares represent terminal nodes. The tree shows characteristics predicting COVID-19 vaccine uptake among Correctional Officers.

At the root node (Layer 1), 68.15% of participants reported taking the COVID-19 vaccine. The most significant determinant of vaccine uptake was respondents’ scores on items measuring general trust in vaccinations (Layer 2). The CHAID algorithm categorized vaccine trust scores into three groups: low (0 – 0.75), moderate (0.76 – 1.33), and high (1.5 – 3). Namely, participants with high trust in vaccines were substantially more likely to report receiving the vaccine.

The second most significant predictor of COVID-19 vaccine uptake was whether participants had ever tested positive for COVID-19 (Layer 3). Taken in tandem, the subgroup most likely to report COVID-19 vaccination included individuals with high trust in vaccines (83.87%) and who had previously tested positive for COVID (Layer 3, Terminal Node 5). Conversely, the subgroup least likely to report vaccination comprised individuals with low trust in vaccines (3.22%) and did not report a previous positive COVID test (0.00%; layer 3, terminal node 2).

### Model 2: Logistic Analysis Results

A subsequent logistic regression analysis was conducted to examine the relationship between vaccine uptake and several predictors, including vaccine beliefs, prior COVID-19 infection (Ever COVID +), age, and education. The results are presented in Table 3.

**Table 3.** Logistic regression of COVID-19 vaccine uptake on vaccine attitudes (n=92) A logistic regression table of COVID-19 vaccine uptake on vaccine attitudes within the study sample size of 92 Correctional Officers.

|  | **Odds Ratio** | **Standard Error** | **z-Score** | **p-Value** |
| --- | --- | --- | --- | --- |
| **Vaccine attitudes** | 1.02** | 0.00 | 3.73 | 0.00 |
| **Ever COVID +** | 1.63 | 1.33 | 1.48 | 0.14 |
| **Age** | 0.59 | 0.26 | -1.25 | 0.21 |
| **Education** | 0.95 | 0.21 | -1.16 | 0.25 |
| **Constant** | 0.03 | 0.04 | -2.45 | 0.01 |

Pseudo $R^2 = 0.29$ ; \*\* $p < 0.01$ , \* $p < 0.05$

Vaccine attitudes emerged as a significant predictor of COVID-19 vaccine uptake. The odds ratio for vaccine attitudes was 1.02 (p < 0.01), indicating that for each one-unit increase in vaccine attitude scores was associated with a 2% increase in the odds of vaccination. A history of COVID-19 infection (Ever COVID +) was positively associated with vaccine uptake (OR= 1.63, p = 0.14), although this relationship was not statistically significant. Age was negatively associated with vaccine uptake (OR= 0.59, p = 0.21), suggesting that older individuals were less likely to be vaccinated, but this effect did not reach statistical significance. Education level was also negatively associated with vaccine uptake (OR= 0.95, p = 0.25), though this relationship was not statistically significant.

## Discussion

Using a novel analysis technique – CHAID modeling – this study demonstrates that COVID-19 vaccine uptake among New England-based COs is most strongly associated with higher levels of vaccine trust and prior, empirically verified exposure to COVID-19. In other words, beliefs that vaccines are safe and necessary, as well as a previous positive COVID-19 test were the most influential factors in whether participants voluntarily received COVID-19 vaccination. Contrary to our hypotheses, COVID-19 vaccine uptake was not associated with other health beliefs, including confidence in evidence-based medical interventions for chronic illness, endorsement of safety-related behaviors, acceptance of behavioral health treatments, or support for public health initiatives. Likewise, vaccine uptake was unrelated to self-reported COVID-19 mitigation behaviors or perceptions of the state’s public health response to COVID-19.

The strongest predictor of COVID-19 vaccination was positive endorsement of the general safety of vaccines, including the Hepatitis B vaccine and the COVID-19 vaccine, as well as the perceived necessity of the COVID-19 vaccine – both its mandate and the belief that the immune system alone is insufficient for maintaining health. Prior studies have identified opposition to vaccine mandates as a primary barrier to vaccine implementation among corrections staff (Kraus et al., 2023), which may partly reflect mistrust of employer-provided vaccine information (Caban-Martinez et al., 2022). Given the centrality of trust in shaping attitudes towards vaccines, it is important not to reduce lack of trust to a simple, monolith category. COs’ opposition to vaccines and mandates may stem from mistrust, distrust, or a combination of both. Trust, mistrust, and distrust are shaped by a combination of empirical and normative inquiry (Jennings et al., 2021) and are socially constructed over time within local contexts that intersect with personal histories (Decoteau & Sweet, 2023; MacDonald, 2015). Mistrust is conceptualized as uncertainty, suggesting the possibility of change, whereas distrust reflects a more static position, a negative assessment of a person, system, idea, or object (Jennings et al., 2021). As such, beliefs about vaccine safety and necessity are relational and individually situated, shaped by trust, mistrust or distrust based on respondents’ lived experiences.

The occupational context of COs may further influence vaccine attitudes. Correctional institutions are structured based on paramilitary organizations, which emphasize hierarchy, discipline, and control (Gilbert, 1997). These norms can foster a culture that devalues vulnerability and stigmatizes behaviors perceived as compliant, such as vaccination, particularly when associated with external mandates. Additionally, correctional environments are predominantly male and often reflect a culture valuing toughness, self-reliance, and skepticism toward perceived weakness (Compton & Brandhorst, 2021). Preventive health behaviors, including vaccination, may therefore be viewed through a gendered lens as unnecessary or “unmanly,” contributing to lower engagement. This is consistent with broader literature suggesting men are less likely to seek healthcare, participate in preventive behaviors, or report illness (Galdas et al., 2005). For COs, where masculine identity and institutional loyalty are intertwined, vaccine hesitancy may be as much about cultural fit and identity as biomedical belief.

Public health interventions within correctional settings must account for these cultural and structural dynamics. Strategies that ignore these factors risk being missed. Messaging delivered by trusted insiders—such as fellow COs, union leaders, or veteran staff—may be more effective than directives from public health agencies or government officials (Caban-Martinez et al., 2022). Framing outreach in terms of strength, duty, and team protection, rather than vulnerability or compliance, may improve engagement and uptake.

This study has several limitations. Participants were drawn from a single New England state, and results should be generalized with caution. While this work contributes to the scant literature on health beliefs of COs, two of the five scales developed for this study exhibited only moderate or acceptable reliability, which may have affected the detection of associations with vaccine uptake. Future research should refine these scales though qualitative methods — such as semi-structure interviews, free listing, or pile sorting —to ensure that items accurately measure their intended constructs. Further investigation is warranted to understand why some scale items performed poorly in this population. In addition to this, the study design unfortunately did not always ensure that the compensation was not provided before the survey administration. This might have encouraged participants to not begin or accurately fill out the survey.

## Conclusion

COs’ willingness to receive the COVID-19 vaccine is critical for protecting incarcerated populations and mitigating the spread of COVID-19 among these high-risk groups, who often have comorbities (Nijhawan, 2016). Despite the widespread impact of COVID-19 in jails and prisons and the proven efficacy of vaccines against hospitalization and death, informational campaigns have not fully addressed distrust and mistrust among correctional staff. Addressing these issues through culturally and occupationally sensitive public health interventions is imperative for improving health outcomes across the criminal-legal system.

## Data Availability

All data produced in the present study are available upon reasonable request to the authors.

## Funding

Dr. Juliette Roddy received funds that supported this research from the Northern Arizona Regional Behavioral Health Authority (NARBHA Institute) James Wurgler, MD Endowed Chair in Criminal Justice and Behavioral Health at Northern Arizona University.

## Acknowledgements

We would like to thank Dr. Bradley Ray for his support with research idea generation. Additionally, we would to thank the Correctional Officers for their time participating in our research.

## Conflict of Interest

The authors have no competing interests to declare. All authors attest they meet the International Committee of Medical Journal Editors (ICMJE) criteria for authorship.

## Notes

### Competing Interest Statement

The authors have declared no competing interest.

### Author Declarations

Health Sciences Institutional Review Board at Tufts Medical Center granted approval for this study in May 2022.

