## Supplemental Material for "An Assessment of Correctional Officers’ Health Beliefs in Relationship to COVID-19 Vaccine Uptake and Hesitancy"

**Supplemental Material: Survey instrument distributed amongst Correctional Officers**

| **SECTION 2: This set of questions asks about your experiences with COVID-19. We ask about serious medical outcomes such as hospitalization and death.** | | |
| --- | --- | --- |
| 1. Outside of work, I have practiced social distancing. | Yes | No |
| 2. Outside of work, I wear personal protective equipment. | Yes | No |
| 3. Since March 2020, I wash my hands more often. | Yes | No |
| 4. Since March 2020, I began to carry hand sanitizer. | Yes | No |
| 5. I agree that the COVID-19 vaccine should be mandated. | Yes | No |
| 6. To prevent the spread of COVID-19, I chose not to see certain family or friends. | Yes | No |
| 7. Governor Baker overstepped his governmental authority in how he responded to COVID-19. | Yes | No |
| 8. Since March 2020, I used an at-home delivery service for groceries or other household goods. | Yes | No |
| 9. Since March 2020, I experienced increased stress. | Yes | No |
| 10. Since March 2020, I have had to pick up more work hours. | Yes | No |
| 11. I have received the COVID-19 vaccine. | Yes | No |
| 12. I have had occasion to be concerned about exposure to someone testing positive for COVID-19. | Yes | No |
| 13. I have been tested for COVID-19. |  |  |
| 14. I have had a positive COVID-19 test. | Yes | No |
| 15. I have been hospitalized for COVID-19. | Yes | No |
| 16. I have personally known someone who had a positive COVID-19 test. | Yes | No |
| 17. I have personally known someone who has died from COVID-19. | Yes | No |

| **SECTION 3: Demographic questions** | | | | | | |
| --- | --- | --- | --- | --- | --- | --- |
| 1. What is the highest degree or level of school you completed? (please circle) | | | | | | |
| 12^th^ Grade – no diploma | | High School Diploma or GED | | | Some College – no degree | |
| Associate degree | | Bachelor’s degree | | | Master’s degree or Above | |
| Other, please describe: | |  | | | | |
| 2. When were you born? (please circle) | | | | | | |
| 1928 – 1945 | 1946 – 1964 | | 1965 – 1980 | 1981 – 1995 | | 1996 – 2010 |

This was the survey instrument that was used in this research study to assess COVID-19-related beliefs and obtain demographic information from the Correctional Officers.
